# Treatment- and population-specific genetic risk factors for anti-drug antibodies against interferon-beta

**DOI:** 10.1101/2020.05.01.20086033

**Authors:** Till F. M. Andlauer, Jenny Link, Dorothea Martin, Malin Ryner, Christina Hermanrud, Verena Grummel, Michael Auer, Harald Hegen, Lilian Aly, Christiane Gasperi, Benjamin Knier, Bertram Müller-Myhsok, Poul Erik Hyldgaard Jensen, Finn Sellebjerg, Ingrid Kockum, Tomas Olsson, Marc Pallardy, Sebastian Spindeldreher, Florian Deisenhammer, Anna Fogdell-Hahn, Bernhard Hemmer, on behalf of the ABIRISK consortium

**Affiliations:** Department of Neurology, Klinikum rechts der Isar, School of Medicine, Technical University of Munich, Ismaninger Str. 22, 81675 Munich, Germany; Max Planck Institute of Psychiatry, Kraepelinstr. 2-10, 80804 Munich, Germany; Department of Clinical Neuroscience, Karolinska Institutet, Visionsgatan 18, 17176 Stockholm, Sweden; Department of Neurology, Medical University of Innsbruck, Anichstr. 35, 6020 Innsbruck, Austria; Institute of Experimental Neuroimmunology, Technical University of Munich, Trogerstr. 9, 81675 Munich, Germany; Institute of Translational Medicine, University of Liverpool, Crown Street, Liverpool, L69 3BX, United Kingdom; Munich Cluster for Systems Neurology (SyNergy), Feodor-Lynen-Str. 17, 81377 Munich, Germany; DMSC, Department of Neurology, Rigshospitalet, University of Copenhagen, 2100 Copenhagen, Denmark; Inflammation, Microbiome and Immunosurveillance, Université Paris-Saclay, INSERM, Faculté de Pharmacie, rue JB Clément, 92290 Châtenay-Malabry, France; Novartis Institutes for Biomedical Research, Novartis Pharma AG, 4056 Basel, Switzerland; Integrated Biologix GmbH, Steinenvorstadt 33, 4051 Basel, Switzerland

**Author notes:** Corresponding authors: Bernhard Hemmer, Till F. M. Andlauer.

**Keywords:** Multiple sclerosis, interferon beta, anti-drug antibodies, human leukocyte antigen (HLA) system, genetics, genome-wide association study, prediction

## Abstract

**Background:** Upon treatment with biopharmaceuticals, the immune system may produce anti-drug antibodies (ADA) that inhibit the therapy. Up to 40% of multiple sclerosis patients treated with interferon β (IFNβ) develop ADA, for which a genetic predisposition exists. Here, we present a genome-wide association study on ADA and predict the occurrence of antibodies in multiple sclerosis patients treated with different interferon β preparations.

**Methods:** We analyzed a large sample of 2,757 genotyped and imputed patients from two cohorts, split between a discovery and a replication dataset. Binding ADA (bADA) levels were measured by capture-ELISA, neutralizing ADA (nADA) titers using a bioassay. Genome-wide association analyses were conducted stratified by cohort and treatment preparation, followed by fixed-effects meta-analysis.

**Results:** Binding ADA levels and nADA titers were correlated and showed a significant heritability (47% and 50%, respectively). The risk factors differed strongly by treatment preparation: The top-associated and replicated variants for nADA presence were the *HLA*-associated variants rs77278603 in IFNβ-1a *s.c.-*(odds ratio (OR)=3.55 (95% confidence interval=2.81-4.48), *p*=2.1×10^−26^) and rs28366299 in IFNβ-1b *s.c*.-treated patients (OR=3.56 (2.69-4.72), *p*=6.6×10^−19^). The rs77278603-correlated *HLA* haplotype *DR15-DQ6* conferred risk specifically for IFNβ-1a *s.c*. (OR=2.88 (2.29-3.61), *p*=7.4×10^−20^) while *DR3-DQ2* was protective (OR=0.37 (0.27-0.52), *p*=3.7×10^−09^). The haplotype *DR4-DQ3* was the major risk haplotype for IFNβ-1b *s.c*. (0R=7.35 (4.33-12.47), *p*=1.5×10^−13^). These haplotypes exhibit large population-specific frequency differences. In a cohort of IFNβ-1a *s.c*.-treated patients, prediction models for nADA reached an AUC of 0.91 (0.85-0.95), a sensitivity of 0.78, and a specificity of 0.90. Patients with the top 30% of genetic risk had, compared to patients in the bottom 30%, an OR of 73.9 (11.8-463.6, *p*=4.4×10^−06^) of developing nADA.

**Conclusions:** We identified several *HLA*-associated genetic risk factors for ADA against interferon β, which were specific for treatment preparations and population backgrounds. Genetic prediction models could robustly identify patients at risk for developing ADA and might be used for personalized therapy recommendations and stratified ADA screening in clinical practice. These analyses serve as a roadmap for genetic characterizations of ADA against other biopharmaceutical compounds.

## Background

Interferon β (IFNβ) preparations are a treatment option for multiple sclerosis (MS). IFNβ-1a is produced in Chinese hamster ovary cells and administered either via intramuscular (*i.m*.) or subcutaneous (*s.c*.) injection. IFNβ-1b is raised using *E. coli* and injected subcutaneously. The amino acid sequence of IFNβ-1b differs at two positions from the mammalian protein [1]. Moreover, IFNβ-1b is not glycosylated, which may affect its immunogenicity, e.g., by promoting the formation of protein aggregates [1, 2]. Posttranslational modifications like deamidation, oxidation, and glycation can also occur spontaneously, depending on the manufacturing and processing of biopharmaceuticals [3]. Therefore, also sequence-identical compounds like IFNβ-1a-*s.c*. and *-i.m*. can differ in their immunogenicity [1, 4].

Up to 40% of patients treated with IFNβ develop anti-drug antibodies (ADA) that bind IFNβ (binding ADA, bADA) [1, 5–7]. A subset of bADA inhibits the interaction of IFNβ with its receptor and thus neutralizes the drug’s biological activity (neutralizing ADA, nADA) [8, 9]. Previous studies have already identified genetic factors influencing the development of ADA but could not establish a consensus on the human leukocyte antigen (*HLA*) alleles [10–17] and single nucleotide polymorphisms (SNPs) [14, 15] contributing to ADA development.

The primary aim of the present study was to characterize the contribution of genetic risk to ADA development by analyzing a large, cross-sectional sample from two different sites: the Karolinska Institutet Stockholm, Sweden (KI), and the Technical University of Munich, Germany (TUM). Both bADA levels and nADA titers were determined in the same patients, allowing for systematic comparisons between the two antibody types. Genome-wide association studies (GWAS) on bADA levels, nADA titers, and nADA presence, as well as analyses of the association of imputed *HLA* alleles with ADA, were conducted. As primary analyses, results were pooled across treatments; as secondary analyses, treatment-specific results were evaluated. The secondary aim of the study was to use these genetic factors for the prediction of ADA development.

## Methods

### Sample inclusion criteria

Patient inclusion criteria were: diagnosis of either clinically isolated syndrome (CIS) or multiple sclerosis (MS); age at first treatment with IFNβ ≥18 years; availability of genotype data and a serum sample fulfilling the sample inclusion criteria. The sample inclusion criteria for bADA-/nADA-negative samples were: ≥12 months of treatment with IFNβ; if more than one sample was eligible, the first sample available at least twelve months after initiation of treatment with IFNβ was selected; no previous positive screening for bADA or nADA. The sample inclusion criteria for previously bADA-/nADA-positive samples were: ≥6 months of treatment with IFNβ; if previously treated with an IFNβ product, not having been ADA-positive during a previous IFNβ treatment period; if more than one sample was eligible, the first sample available at least six months after initiation of treatment with IFNβ was selected. Based on these criteria, 1,810 patients were eligible at KI and 1,488 at TUM. The respective local ethics committees approved the study, and all participants provided written informed consent.

### Power calculation

In a previous ADA GWAS, Weber *et al*. identified a genome-wide significant SNP explaining 2.5% of the variance of bADA levels [14]. To have sufficient power for identification of additional associated variants, 2,000 patients were assigned to the discovery-stage analyses. In this dataset, 80% of statistical power can be reached for a variant explaining 1.96% of the variance at a *p*-value of 5×10^−8^ (calculated using the *R* package *pwr*). Effect sizes in the replication stage are expected to be smaller than in the discovery stage [18]. We thus estimated that at least 682 patients are necessary for replicating up to ten linkage disequilibrium (LD)-independent signals with 80% power, explaining 1.7% of the variance using a one-sided hypothesis. Because of an expected reduction in power due to heterogeneity and an expected decrease in the number of available samples after titration and quality control (QC), we initially selected 800 patients for the replication stage.

### Selection of patients

To select approximately 2,800 patients for ADA screening and titration, all available previously bADA-/nADA-positive samples (n=984) were combined with previously ADA-negative samples (n=2,314) best-matching ADA-positive ones [Additional file 1]. Propensity score matching was conducted using the *R* package *optmatch* [19], based on recruitment site, gender, the age at the blood draw, the IFNβ treatment preparation, the total duration of IFNβ treatment, and eight multidimensional scaling (MDS) ancestry components of the genetic identity-by-state (IBS) matrix, calculated from the genotype data to account for population stratification [Additional file 2]. From the selected patients, new bADA levels and nADA titers (see below) could be determined for 938 previously bADA-/nADA-positive and 1,819 previously ADA-negative samples (Table 1 and [Additional file 3]). These patients were randomized into a discovery (n=2,000), and a replication (n=757) set, using adaptive randomization to minimize differences regarding recruitment site, nADA measurement site (Innsbruck or Copenhagen, see below), gender, the age at the blood draw, the IFNβ treatment preparation, and the total duration of IFNβ treatment.

**Table 1:** Sample characteristics **Table 1:** N (%) refers to the entire cohort, the other percentages to the respective column. The nADA and bADA measurements shown here were obtained within the present study. Non-parametric summary statistics are provided for variables that were not normally distributed. Progressive MS = patients with a primary or secondary progressive disease course, as opposed to clinically isolated syndrome and relapsing-remitting MS. KI = Karolinska Institutet, Sweden; TUM = Technical University of Munich, Germany; SD = standard deviation; MAD = median absolute deviation.

| Treatment preparation | IFNβ-1a *i.m.* | | IFNβ-1a *s.c.* | | IFNβ-1b *s.c.* | |
| --- | --- | --- | --- | --- | --- | --- |
| Cohort | KI Sweden | TUM Germany | KI Sweden | TUM Germany | KI Sweden | TUM Germany |
| N (%) | 345 (24.7) | 251 (18.4) | 590 (42.3) | 558 (40.9) | 459 (32.9) | 554 (40.6) |
| Mean age (SD) | 46.7 (9.91) | 40.1 (9.61) | 44.1 (9.9) | 38.9 (9.62) | 45.4 (10.4) | 41.4 (10.4) |
| Female sex (%) | 216 (62.6) | 191 (76.1) | 440 (74.6) | 406 (72.8) | 334 (72.8) | 390 (70.4) |
| Median treatment duration in months (MAD) | 21.0 (8.05) | 40.0 (20.4) | 30.0 (15) | 55.2 (23.1) | 24.9 (12.9) | 46.9 (23.4) |
| Progressive MS (%) | 60 (17.4) | 34 (13.5) | 120 (20.3) | 90 (16.1) | 130 (28.3) | 128 (23.1) |
| nADA positive (%) | 45 (13.0) | 41 (16.3) | 204 (34.6) | 188 (33.7) | 245 (53.4) | 255 (46) |
| Median nADA titer (MAD)<br>*nADA-positive samples* | 320 (280) | 320 (280) | 640 (600) | 1280 (1240) | 320 (280) | 320 (280) |
| Median bADA level (MAD)<br>*all samples* | 13.9 (6.5) | 9.6 (5.7) | 23.2 (13.9) | 16.3 (10.5) | 35.8 (19.9) | 29.4 (20.0) |
| Median bADA level (MAD)<br>*nADA-positive samples* | 63.8 (42.6) | 25.8 (22.3) | 109.0 (84.8) | 115.0 (104.0) | 69.0 (37.0) | 73.8 (50.7) |

### ADA screening and titration

Binding ADA levels were measured by capture ELISA [20] at a single site (Munich) and were calculated from optical densities using a standard curve [Additional file 2]. For the assessment of nADA titers, measured as the inverse of serum dilutions using a luciferase-based bioassay [21], samples were first screened, and titration was only conducted for samples positive during screening [22]. Assessment of nADA titers was conducted at two separate sites (Innsbruck and Copenhagen), to which samples were assigned using adaptive randomization to minimize differences regarding the recruitment site, gender, the age at the blood draw, the IFNβ treatment preparation, and the total duration of IFNβ treatment. We obtained 2,748 valid measurements for nADA screening and titers as well as 2,752 bADA levels; for 2,743 patients, both nADA titers and bADA levels were available (1,990 in the discovery and 753 in the replication set). The presence of nADA was defined as samples positive in the screening for nADA and showing a nADA titer >0. Correlations of bADA and nADA were calculated in a combined dataset of all samples. For the estimation of the nADA status from bADA levels, the cutoff was established using nested cross-validation in the discovery dataset [Additional file 2]. Sensitivity and specificity were calculated by the application of this cutoff to the replication data.

### Genotyping and imputation

SNPs were genotyped on Illumina microarrays, and QC was conducted separately for KI and TUM data in PLINK v1.90b3.44 or higher [23], as described before [24]. Genotype data were imputed to the 1000 Genomes Phase 3 reference panel using SHAPEIT2 and IMPUTE2 [25–27]. The resulting datasets contained 9,096,778 and 8,550,834 high-quality variants with a MAF≥1% for KI and TUM, respectively. *HLA* allele imputation was performed using SNP2HLA v1.0.3 / Beagle v3.04 and the Type 1 Diabetes Genetics Consortium imputation panel, as previously described [28–30]. The extended haplotypes were determined based on the haplotype phasing estimated in Beagle. An additional file provides further details on QC and imputation [Additional file 2].

### Estimation of heritability and GWAS

ADA titers/levels were transformed by rank-based inverse normal transformation before analyses. Sex, age, treatment preparation, treatment duration, titration site, and eight ancestry components were used as covariates in all analyses. The covariate treatment preparation was also used in preparation-specific analyses and controlled, beyond the three preparation types, for: a), whether treatment with IFNβ-1a *s.c*. had begun before 2008 (change of the formulation [31]), b), in the TUM cohort, the dose of IFNβ-1a *s.c*. (22 vs. 44 μg), c), in the KI cohort, the IFNβ-1b *s.c*. brand used.

The SNP heritability and genetic correlations were estimated with GCTA GREML on a combined dataset of KI and TUM genotypes [32–35], using the covariates mentioned above plus treatment preparation and the recruitment site.

For GWAS, ADA titers/levels were analyzed by linear regression models, the presence of nADA by logistic regression. Samples from Sweden and Germany were analyzed separately in PLINK; results were combined using fixed-effects metaanalysis in METAL [36]. For plots of the ancestry components in both cohorts, see [Additional file 4]. Analyses were run stratified by treatment (IFNβ-1a *i.m*., IFNβ-1a *s.c*., IFNβ-1b *s.c*.), and the groups were combined via fixed-effects metaanalysis after pooling the two cohorts. Additional details are provided [Additional file 2].

The threshold for genome-wide significance was α=5×10^−8^. For replication, the significance threshold α was corrected for the total number of variants analyzed across all SNP-based analyses in the replication phase (n=16) using Bonferroni’s method, *i.e*., α=0.05/16=3×10^−3^.

### Permutation analyses

All replicated associations from hypothesis-free linear regression analyses were validated using permutation analyses. In these analyses, the null distribution of test statistics was empirically determined by repeating regression analyses either 200 or 1 million times with random sampling of phenotype data. To calculate a *p*-value, the number of tests were counted where a model with a random genotype-phenotype association showed the same or a more extreme *p*-value than the correct, non-randomized mode; this number was divided by the total number of tests (200 or 1 million). Permutation-based *p*-values were pooled per cohort and treatment using Stouffer’s Z-score method [37]. For GWAS variants, 200 million permutations per dataset (discovery / replication), cohort, and treatment preparation were carried out (allowing for *p*-values down to 1×10^−8^), for stepwise conditional models and *HLA* alleles, the default was 1 million permutations per group (for *p*-values down to 1×10^−6^). If these permutation *p*-values were <1×10^−6^, 200 million permutations were conducted. If the permutation *p*-values were <1×10^−8^, they were set to 1×10^−8^.

### EQTL analyses

The significant *cis*-expression quantitative trait loci (eQTLs) in whole blood were looked up in the GTEx v8 database (https://gtexportal.org/) downloaded on April 1^st^, 2020 (dbGaP accession number phs000424.v8.p2) [38].

### Gene-set analyses

Gene-set analyses were conducted with MAGMA v1.07b [39]. First, SNPs within gene boundaries were annotated to RefSeq genes (0 bp window). Second, gene analysis was performed on the pooled GWAS summary statistics, based on LD information from the 1000 Genomes EUR reference panel, using both mean- and top-SNP gene models. Third, gene-level analyses used a combination of the curated 186 KEGG and 1499 Reactome pathways from the MSigDB 7.0 database gene sets [40].

### *HLA* and stepwise conditional analyses

The association of *HLA* alleles was analyzed in *R* v3.3 or higher. As in the GWAS, sex, age, treatment preparation and duration, titration site, and eight ancestry components were used as covariates. Separate regression models were run per cohort and treatment preparation, followed by a two-level meta-analysis: results were combined using fixed-effects meta-analysis first by cohort and then by preparation. For assessment of significance, we applied Bonferroni correction for testing 131 alleles and extended haplotypes [41] (rounded down to α=3×10^−4^). In the replication phase, we corrected for multiple testing of 41 *HLA* alleles and haplotypes prioritized across all analyses (rounded down to α=1×10^−3^). Note that the associations of all *HLA* alleles and haplotypes presented in this study also reached genome-wide significance (*p*<5×10^8^) in the pooled analyses of discovery and replication samples, except for the super-extended haplotypes *C7-DQ6* and *A3-DQ6*, which reached a *p*<10×10^−8^.

Stepwise conditional regression was conducted, as previously described [42, 43], first only for *HLA* alleles and then for a joint dataset of *HLA* alleles and SNPs mapping to the extended MHC region. In brief, the association of all alleles/SNPs was first tested in separate regression models. The top-associated allele/SNP was then added as a covariate to the regression model, and the analysis was repeated for all remaining alleles/SNPs. This addition of top-associated alleles/SNPs as covariates was repeated until no allele/SNP was significant anymore after correction for multiple testing.

### Polygenic risk scores (PRS) and prediction of nADA

PRS were calculated in *R* v3.33 using imput In each outer cross-validation instance, the cutoff producing the maximum sensitivity across three inner cross-validation folds was tested on the remaining fold. Nested cross-validation was repeated 100 times and the mean cutoff of the 100 repetitions was used as the final cutoff.ed genetic data, as described previously [44, 45]. For each PRS, the effect sizes of variants from the discovery-stage analyses (training data), below a selected discovery-stage *p*-value threshold, were multiplied by the imputed SNP dosage in the replication-stage test data and then summed to produce a single PRS per threshold. For each analysis group, eight PRS based on different GWAS *p*-value thresholds were calculated on the discovery data. Additional details are provided [Additional file 2].

For the prediction of the presence of nADA in the replication dataset, logistic regression of the eight PRS, the top single GWAS variant, and the top *HLA* allele from the discovery stage was conducted using the GWAS models. The area under the receiver operating characteristic curve (AUC) was calculated using the *R* package *pROC*, its 95 % confidence interval (CI) with the function *ci.auc* (2000 stratified bootstrap replicates). At this stage, we adapted the significance threshold for ten tests using Bonferroni correction (Fig 3a-b). For each treatment preparation, the model with the highest AUC was selected. The performance of all top models was subsequently compared, with a significance threshold adapted for 160 comparisons (α=3.13×10^−4^, Fig 3c).

The sensitivity and specificity of the predictions were calculated using the package *OptimalCutpoints*, maximizing both measures (*MaxSpSe*). To avoid overfitting, the cutpoint was selected using nested cross-validation with three outer and four inner folds. In each outer cross-validation instance, the cutoff producing the maximum sensitivity across three inner cross-validation folds was tested on the remaining fold. Nested cross-validation was repeated 100 times and the mean cutoff of the 100 repetitions was used as the final cutoff. Nagelkerke’s pseudo-*R*^2^ was calculated using the package *fmsb*. For a comparison of patients either at low or high genetic risk, patients within the lower 30% of genetic risk were compared to the patients in the upper 30%. We initially selected a 10% cutoff for this contrast and increased it in 10% steps until the sample size in the replication dataset sufficed for the stable convergence of regression models.

## Results

From 2,757 MS patients recruited in Sweden and Germany and treated with three different IFNβ preparations (Table 1), bADA levels were measured by capture ELISA [20] and nADA titers using a luciferase-based bioassay [21, 22] [Additional files 1-3]. The bADA levels were correlated with nADA presence (Spearman *ρ*=0.66) and nADA titers (*ρ*=0.71). Compared to the presence of nADA determined via screening and titration, estimation of the nADA status from bADA levels had a sensitivity=0.85 and a specificity=0.84 (Fig 1).

**Fig. 1:**
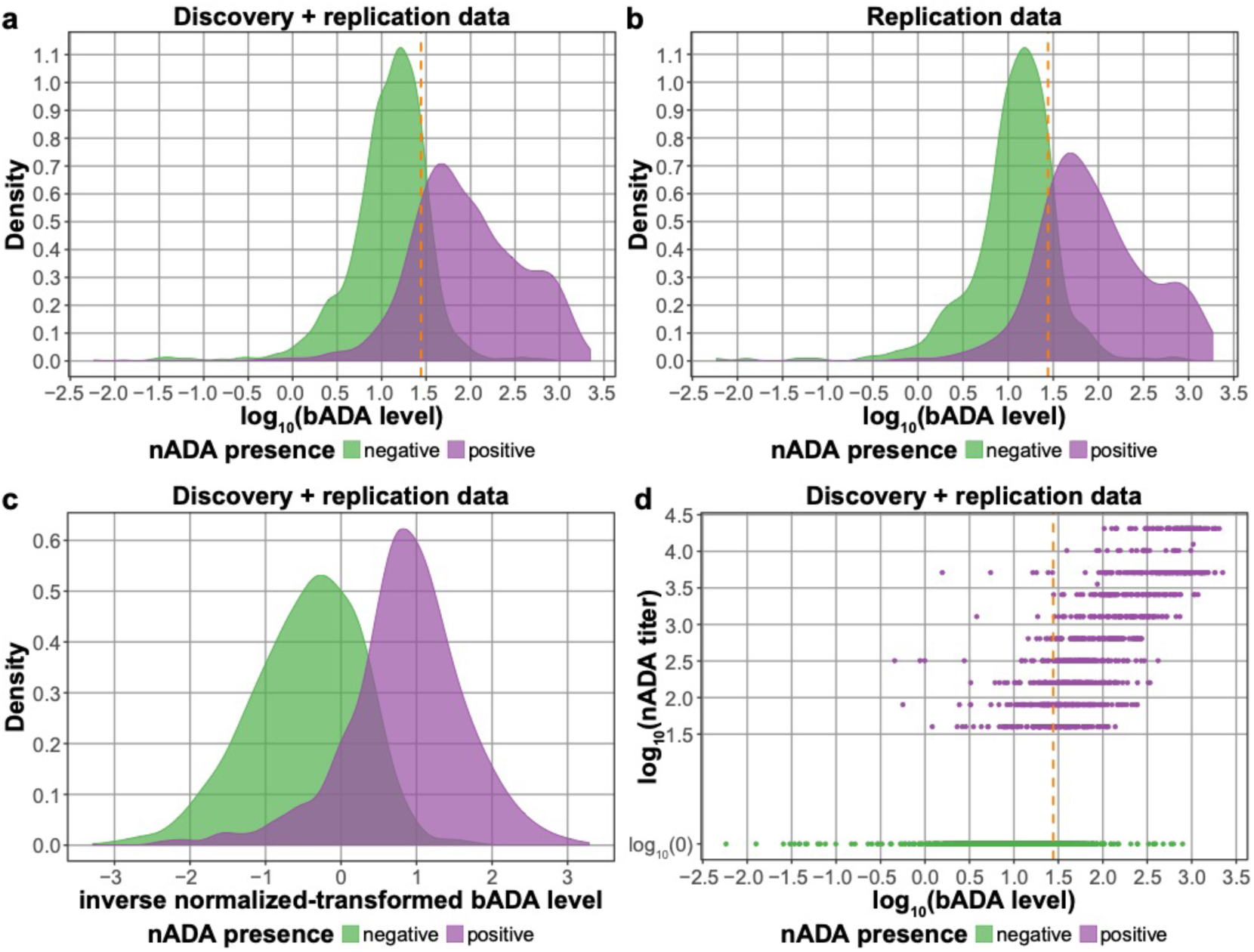
Comparison of bADA levels and nADA titers. The orange dashed line indicates the cutoff at a log_10_ bADA level of 1.442256, which optimized the maximum sensitivity and specificity in the discovery data. This cutoff had a sensitivity = 0.83 and a specificity = 0.82 in the discovery dataset; a sensitivity = 0.85 and a specificity = 0.84 in the replication dataset; and a sensitivity = 0.84 and a specificity = 0.82 in the combined dataset. **a** Density plot showing log_10_ bADA levels in the combined discovery and replication dataset stratified by nADA presence. **b** Density plot showing log_10_ bADA levels in the replication data stratified by nADA presence. **c** Density plot showing rank-based inverse-normal transformed bADA levels in the combined dataset stratified by nADA presence. **d** Comparison of log_10_ bADA levels to log_10_ nADA titers in the combined dataset, colored by nADA presence.

### SNP heritability and genetic correlations

The SNP-based heritability estimated from the genotype data was 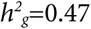 (standard error (SE)=0.15, *p*=1.4×10^−4^) for the inverse-normal transformed bADA levels and 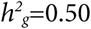 (SE=0.15, *p*=2.9×10^−4^) for the transformed nADA titers. The SNP heritability of the presence of nADA was 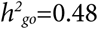 on the observed scale (SE=0.15, *p*=4.9×10^−4^) and, assuming an incidence of 0.35 for ADA, 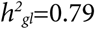 (SE=0.25) on a liability scale. Genetic correlations of bADA levels with nADA presence (*r_g_*=0.89, SE=0.14, *p*=1.2×10^−3^) and titers (*r_g_*=0.95, SE=0.11, *p*=7.0×10^−4^) were very high. The nADA titers in the subset of nADA-positive patients did not show a significant SNP heritability (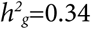, SE=0.44, *p*=0.22) and was thus not analyzed further.

### GWAS of nADA presence across treatment preparations

For all further analyses, the patients were randomized into a discovery (n=2,000), and a replication (n=757) set [Additional file 3]. In the GWAS pooled for the three treatment preparation groups, only variants within the major histocompatibility complex (MHC) region were significant on a genome-wide scale (*p*<5×10^−8^) in the discovery-stage analyses and replicated [Additional files 5 and 6]. There was no indication for systematic inflation of test statistics; the genomic inflation factors λ were in the expected range [Additional file 7].

In the discovery GWAS of nADA presence, analyzed by logistic regression, the strongest association was observed for the insertion TTTTTTT of the variant rs9281971 (Table 2 and [Additional files 8 and 9]). This insertion had a frequency of 36.0% in Swedish and 38.6% in German patients. No other genome-wide significant variant with linkage disequilibrium (LD) *r*^2^<0.2 with the top signal was identified. The insertion replicated at genome-wide significance and was also the top association signal in a pooled analysis of discovery and replication data (discovery: odds ratio (0R)=0.59 (95% confidence interval (CI)=0.50-0.69), *p*=1.9×10^−11^; replication: OR=0.44 (0.32-0.59), *p_one-sided_=*2.4×10^−08^; discovery + replication: OR=0.55 (0.48-0.63), *p*=2.3×10^−17^). Inversely, for each copy lacking the insertion TTTTTTT at this site, the OR associated with risk for nADA was thus 1.82 (1.58-2.08). This association was supported in all three treatment groups but was strongest in IFNβ-1a *s.c*.-treated patients [Additional file 10]. Because the MHC region shows long-range LD patterns, we conducted stepwise conditional regression analyses in the pooled dataset (discovery + replication) as a second approach to estimate the number of independently associated signals. No variant except rs9281971-TTTTTTT was significantly associated with nADA presence on a genome-wide scale [Additional file 11].

**Table 2:** Genome-wide significant variants from GWAS analyses. **Table 2:** The top association signals within the MHC region that showed genome-wide significance in the discovery-stage GWAS (α=5×10^−8^) and replicated (α=3×10^−3^). (**A**) Results in a pooled analysis of all three treatment preparations. (**B-C**) Results for (**B**) IFNβ-1a *s.c*.-treated and (**C**) IFNβ-1b *s.c*.-treated patients. For additional details, see [Additional file 8]. Chr. = chromosome; Pos. = position in base pairs (build *hg19);* MA = minor and effect allele; MAF = minor allele frequency; KI = Karolinska Institutet, Sweden; TUM = Technical University of Munich, Germany; β = effect size; OR = odds ratio; pres. = presence; (T)_7_ = TTTTTTT.

| ADA | Variant | Chr. | Pos. (bp) | MA | MAF |  | Discovery |  | Replication |  | Pooled |  |
| --- | --- | --- | --- | --- | --- | --- | --- | --- | --- | --- | --- | --- |
| **A: All preparations** |  |  |  |  | KI | TUM | OR | *p* | OR | *p* (one-sided) | OR | *p* |
| nADA pres. | rs9281971 | 6 | 32596722 | (T)<sub>7</sub> | 0.36 | 0.39 | 0.59 | 1.9×10<sup>-11</sup> | 0.44 | 2.4×10<sup>-08</sup> | 0.55 | 2.3×10<sup>-17</sup> |
|  |  |  |  |  |  |  | β |  | β |  | β |  |
| nADA titer | rs9271377 | 6 | 32587165 | G | 0.30 | 0.33 | -0.15 | 4.0×10<sup>-11</sup> | -0.17 | 4.0×10<sup>-05</sup> | -0.16 | 1.5×10<sup>-14</sup> |
| nADA titer | rs9281971 | 6 | 32596722 | (T)<sub>7</sub> | 0.36 | 0.39 | -0.14 | 1.4×10<sup>-10</sup> | -0.23 | 4.6×10<sup>-09</sup> | -0.16 | 2.5×10<sup>-17</sup> |
| bADA level | rs9271377 | 6 | 32587165 | G | 0.30 | 0.33 | -0.23 | 1.3×10<sup>-13</sup> | -0.20 | 8.5×10<sup>-05</sup> | -0.22 | 1.2×10<sup>-16</sup> |
| bADA level | rs9272071 | 6 | 32599487 | C | 0.32 | 0.38 | -0.21 | 1.8×10<sup>-11</sup> | -0.28 | 2.0×10<sup>-08</sup> | -0.23 | 4.5×10<sup>-18</sup> |
| **B: IFNβ-1a s.c.** |  |  |  |  |  |  | OR |  | OR |  | OR |  |
| nADA pres. | rs77278603 | 6 | 32469421 | A | 0.43 | 0.40 | 3.33 | 5.3×10<sup>-19</sup> | 4.43 | 1.9×10<sup>-09</sup> | 3.55 | 2.1×10<sup>-26</sup> |
| nADA pres. | rs9271700 | 6 | 32593198 | G | 0.42 | 0.39 | 3.16 | 8.6×10<sup>-19</sup> | 5.08 | 1.2×10<sup>-10</sup> | 3.48 | 5.4×10<sup>-27</sup> |
|  |  |  |  |  |  |  | β |  | β |  | β |  |
| nADA titer | rs77278603 | 6 | 32469421 | A | 0.43 | 0.40 | 0.38 | 2.2×10<sup>-19</sup> | 0.36 | 8.1×10<sup>-10</sup> | 0.37 | 2.4×10<sup>-27</sup> |
| nADA titer | rs9271673 | 6 | 32592833 | C | 0.41 | 0.39 | 0.37 | 3.0×10<sup>-19</sup> | 0.38 | 5.2×10<sup>-11</sup> | 0.37 | 2.1×10<sup>-28</sup> |
| bADA level | rs9281962 | 6 | 32594597 | T | 0.44 | 0.43 | 0.51 | 6.0×10<sup>-22</sup> | 0.51 | 5.5×10<sup>-12</sup> | 0.51 | 4.6×10<sup>-32</sup> |
| **C: IFNβ-1b s.c.** |  |  |  |  |  |  | OR |  | OR |  | OR |  |
| nADA pres. | rs28366299 | 6 | 32560870 | A | 0.20 | 0.19 | 3.11 | 2.1×10<sup>-12</sup> | 5.84 | 5.2×10<sup>-09</sup> | 3.56 | 6.6×10<sup>-19</sup> |
|  |  |  |  |  |  |  | β |  | β |  | β |  |
| nADA titer | rs28366299 | 6 | 32560870 | A | 0.20 | 0.19 | 0.40 | 1.8×10<sup>-16</sup> | 0.52 | 1.1×10<sup>-09</sup> | 0.43 | 5.3×10<sup>-24</sup> |
| nADA titer | rs9272775 | 6 | 32610257 | C | 0.22 | 0.23 | 0.38 | 2.5×10<sup>-16</sup> | 0.51 | 3.2×10<sup>-10</sup> | 0.41 | 2.5×10<sup>-24</sup> |
| bADA level | rs78279385 | 6 | 32451758 | A | 0.23 | 0.25 | 0.39 | 2.7×10<sup>-14</sup> | 0.39 | 2.0×10<sup>-06</sup> | 0.39 | 5.5×10<sup>-19</sup> |
| bADA level | rs9272775 | 6 | 32610257 | C | 0.22 | 0.23 | 0.39 | 1.0×10<sup>-13</sup> | 0.46 | 1.1×10<sup>-07</sup> | 0.41 | 1.6×10<sup>-19</sup> |

### GWAS of bADA levels and ADA titers across treatment preparations

For both bADA levels and nADA titers, analyzed by linear regression, the top-associated SNP in the discovery GWAS was rs9271377, which replicated for both measures (Table 2). The SNP is correlated with nearby (9,558 base pairs (bp) downstream) rs9281971-TTTTTTT (LD *r*^2^=0.39 in Swedish and *r*^2^=0.51 in German patients). In the pooled analysis of discovery and replication data, this rs9281971-TTTTTTT was the strongest association for nADA titers and the nearby (2,759 bp downstream), highly correlated (*r*^2^=0.77 in Swedish and *r*^2^=0.81 in German patients) SNP rs9272071 for bADA levels. These associations were supported in all three treatment groups, but to a lesser degree in IFNβ-1a *i.m*.-treated patients [Additional file 10]. Because of possible deviations from normality, the associations of all replicated ADA variants were confirmed using nonparametric permutation analyses [Additional file 8]. In stepwise conditional regression analyses in the pooled dataset, rs9281971-TTTTTTT was the only significant variant for nADA titers, while four variants reached significance for bADA levels (rs9272071, rs28746882, rs1265086, and *HLA-DRB1*04:04* [Additional file 11]).

All three top-associated variants map directly upstream of the gene *HLA-DQA1* (rs9271377 18.0 kbp, rs9281971 8.5 kbp, and rs9272071 5.7 kbp upstream [Additional file 9]). The variants were all in weak to moderate LD with the *HLA* allele *DQA1 *05:01* (LD range: 0.31≥*r*^2^≤0.50) and part of cis-expression quantitative trait loci (eQTLs) with *HLA-DRB5* (GTEx v8 [Additional file 12]) [38]. In gene-set analyses using KEGG and Reactome gene sets [39, 40], several immune-related pathways were significant after correction for multiple testing, e.g., “antigen processing and presentation”, “Translocation of ZAP-70 to Immunological synapse”, and “PD-1 signaling” [Additional file 13].

### Analysis of *HLA* variants across treatment preparations

Next, we analyzed the association of imputed *HLA* alleles with ADA (Table 3). The full list of significantly associated *HLA* alleles and haplotypes is shown in [Additional file 14]. In analyses of nADA presence and titers across all treatment preparations, no *HLA* allele was significant after correction for multiple testing and replicated.

**Table 3:** Selected significant *HLA* alleles and haplotypes. **Table 3:** Selected four-digit *HLA* alleles and extended haplotypes that were significantly associated (*p*<3×10^−4^) with an ADA measurement and replicated (*p_one-sided_*<1 ×10^−3^). Alleles that are part of one of the listed extended haplotypes and showed a similar or weaker association than the haplotypes and which did not remain significant when conditioning for the haplotypes are not displayed separately; for a full table of all results see [Additional file 14]. HT = haplotype; AF = allele frequency; KI = Karolinska Institutet, Sweden; TUM = Technical University of Munich, Germany; β = effect size; OR = odds ratio; pres. = presence.

| ADA | Allele / HT | AF |  | Discovery |  | Replication |  | Pooled |  |
| --- | --- | --- | --- | --- | --- | --- | --- | --- | --- |
| A: Risk alleles (all preparations) | | KI | TUM | $\beta$ | $p$ | $\beta$ | $p_{one-sided}$ | $\beta$ | $p$ |
| bADA levels | HLA-DQA1*01:02 | 0.42 | 0.36 | 0.18 | $1.6 \times 10^{-08}$ | 0.17 | $3.7 \times 10^{-04}$ | 0.17 | $4.8 \times 10^{-11}$ |
| bADA levels | B7-DQ6 | 0.18 | 0.17 | 0.17 | $8.6 \times 10^{-06}$ | 0.21 | $3.8 \times 10^{-04}$ | 0.18 | $2.7 \times 10^{-08}$ |
| B: Protective alleles (all preparations) |  |  |  |  |  |  |  |  |  |
| bADA levels | DR3-DQ2 | 0.13 | 0.12 | -0.23 | $9.6 \times 10^{-08}$ | -0.21 | $6.7 \times 10^{-04}$ | -0.23 | $5.0 \times 10^{-10}$ |
| C: Risk alleles (IFNβ-1a s.c.) | | | | OR | | OR | | OR | |
| nADA pres. | DR15-DQ6 | 0.34 | 0.30 | 2.73 | $3.1 \times 10^{-14}$ | 3.41 | $1.5 \times 10^{-07}$ | 2.89 | $7.4 \times 10^{-20}$ |
| | | | | $\beta$ | | $\beta$ | | $\beta$ | |
| nADA titer | DR15-DQ6 | 0.34 | 0.30 | 0.36 | $1.3 \times 10^{-15}$ | 0.32 | $2.3 \times 10^{-07}$ | 0.35 | $3.6 \times 10^{-21}$ |
| bADA levels | DR15-DQ6 | 0.34 | 0.30 | 0.44 | $6.8 \times 10^{-15}$ | 0.45 | $4.4 \times 10^{-08}$ | 0.44 | $3.5 \times 10^{-21}$ |
| D: Protective alleles (IFNβ-1a s.c.) | | | | OR | | OR | | OR | |
| nADA pres. | DR3-DQ2 | 0.13 | 0.12 | 0.40 | $9.1 \times 10^{-07}$ | 0.29 | $4.0 \times 10^{-04}$ | 0.37 | $3.7 \times 10^{-09}$ |
| | | | | $\beta$ | | $\beta$ | | $\beta$ | |
| nADA titer | DR3-DQ2 | 0.13 | 0.12 | -0.31 | $8.6 \times 10^{-08}$ | -0.28 | $5.0 \times 10^{-04}$ | -0.30 | $3.4 \times 10^{-10}$ |
| bADA levels | DR3-DQ2 | 0.13 | 0.12 | -0.41 | $2.1 \times 10^{-08}$ | -0.40 | $1.5 \times 10^{-04}$ | -0.41 | $2.5 \times 10^{-11}$ |
| E: Risk alleles (IFNβ-1b s.c.) | | | | OR | | OR | | OR | |
| nADA pres. | HLA-DRB1*04:01 | 0.09 | 0.07 | 6.82 | $3.4 \times 10^{-13}$ | 14.7 | $1.7 \times 10^{-07}$ | 7.95 | $1.4 \times 10^{-18}$ |
| nADA pres. | DR4-DQ3 | 0.07 | 0.05 | 6.23 | $1.2 \times 10^{-09}$ | 14.7 | $6.1 \times 10^{-06}$ | 7.35 | $1.5 \times 10^{-13}$ |
| | | | | $\beta$ | | $\beta$ | | $\beta$ | |
| nADA titer | HLA-DRB1*04:01 | 0.09 | 0.07 | 0.56 | $4.1 \times 10^{-15}$ | 0.56 | $9.2 \times 10^{-06}$ | 0.56 | $3.7 \times 10^{-19}$ |
| nADA titer | DR4-DQ3 | 0.07 | 0.05 | 0.50 | $1.6 \times 10^{-09}$ | 0.54 | $1.3 \times 10^{-04}$ | 0.51 | $1.9 \times 10^{-12}$ |
| bADA levels | HLA-DRB1*04:01 | 0.09 | 0.07 | 0.62 | $4.6 \times 10^{-15}$ | 0.61 | $1.8 \times 10^{-06}$ | 0.62 | $8.7 \times 10^{-20}$ |
| bADA levels | DR4-DQ3 | 0.07 | 0.05 | 0.56 | $4.9 \times 10^{-10}$ | 0.53 | $2.2 \times 10^{-04}$ | 0.56 | $9.0 \times 10^{-13}$ |

For bADA levels, the top-associated and replicated *HLA* risk allele across preparations was *HLA-DQA1*01:02* (*p*=4.79×10^−11^). This allele is part of the extended ancestral haplotype *B7-DQ6* (the combined presence of *HLA-B*07:02, HLA-DRB1*15:01*, *HLA-DQA1*01:02*, and *HLA-DQB1*06:02* on the same chromosome), which was also associated (*p*=2.70×10^−08^). However, conditional analyses indicated an independent effect of *HLA-DQA1*01:02* from the extended haplotype [Additional file 14]. The extended ancestral haplotype *DR3-DQ2* (the combined presence of *HLA-DRB1 *03:01*, *HLA-DQA1*05:01*, and *HLA-DQB1*02:01* on the same chromosome) was the top protective association for bADA levels across preparations (*p*=4.97×10^−10^; Table 3 [Additional file 14]). In conditional analyses, none of these *HLA* associations were independent of the top bADA-SNP rs9271377 [Additional file 14].

### Treatment-specific *HLA* analyses

For many identified *HLA* alleles, support for an association was predominantly observed in patients treated with either IFNβ-1a *s.c*. or -1b *s.c*. but not in both groups simultaneously [Additional file 10]. We, therefore, also conducted analyses separately for the two main treatment preparations used in our cohorts (IFNβ-1a *s.c*. and IFNβ-1b *s.c*.). We did not conduct hypothesis-free preparation-specific analyses for IFNβ-1a *i.m*., with its much smaller number of ADA-positive patients (Table 1).

For all three ADA measurements (bADA levels, nADA titers, and nADA presence), allele *HLA*-*DQB1*06:02* and the ancestral haplotype *DR15-DQ6*, both smaller subsets of *B7-DQ6*, were the risk alleles showing the most robust support for an association in IFNβ-1a *s.c*.-treated patients (*DR15-DQ6* nADA presence: OR=2.88 (2.29-3.61), *p*=7.4×10^−20^; Fig 2a, Table 3 [Additional file 14]). All other risk alleles associated with IFNβ-1a *s.c*.-induced ADA were part of this extended haplotype and did not remain significant when conditioning for *DR15-DQ6*. None of these variants passed the discovery-stage significance threshold in patients receiving IFNβ-1b *s.c*. [Additional file 14].

Similarly, the ancestral haplotype *DR3-DQ2* and its allele *HLA-DQB1 *02:01* were the protective alleles showing the lowest *p*-values in IFNβ-1a *s.c*.-treated patients (*DR3-DQ2* nADA presence: OR=0.37 (0.27-0.52), *p*=3.7×10^−9^; Table 3 [Additional file 14]), with all other protective alleles being part of this extended haplotype. No other allele remained significant when conditioning for *DR3-DQ2*. None of these variants were significantly associated in patients treated with IFNβ-1b *s.c*. (e.g., *DR3-DQ2*, nADA presence *p*=0.27) or IFNβ-1a *i.m*. after correction for multiple testing [Additional file 14].

In IFNβ-1b *s.c*.-treated patients, *HLA-DRB1*04:01* was the risk allele that showed the most robust support for an association for all three ADA measurements, and all associated alleles were part of the haplotype *DR4-DQ3* (the combined presence of *HLA-DRB1*04:01, HLA-DQA1*03:01*, and *HLADQB1*03:02* on the same chromosome). The pooled association strength of *DR4-DQ3* for nADA presence was OR=7.35 (4.32-12.47), *p*=1.5×10^−13^ in patients treated with IFNβ-1b *s.c*. (Fig 2b, Table 3 [Additional file 14]). Of note, when conditioning for *DR4-DQ3*, the association of *HLA-DRB1*04:01* remained significant, suggesting it to constitute the primary signal [Additional file 14]. However, statistical power for fine-mapping this signal was limited because of the low allele frequencies of the allele and the haplotype (Table 3). These alleles and the haplotype were not significantly associated in IFNβ-1a *s.c*.-treated patients (e.g., *DR4-DQ3*, nADA presence for IFNβ-1a *s.c., p*=0.22 [Additional file 14]). There were no significant protective alleles for patients receiving IFNβ-1b *s.c*.

**Fig. 2:**
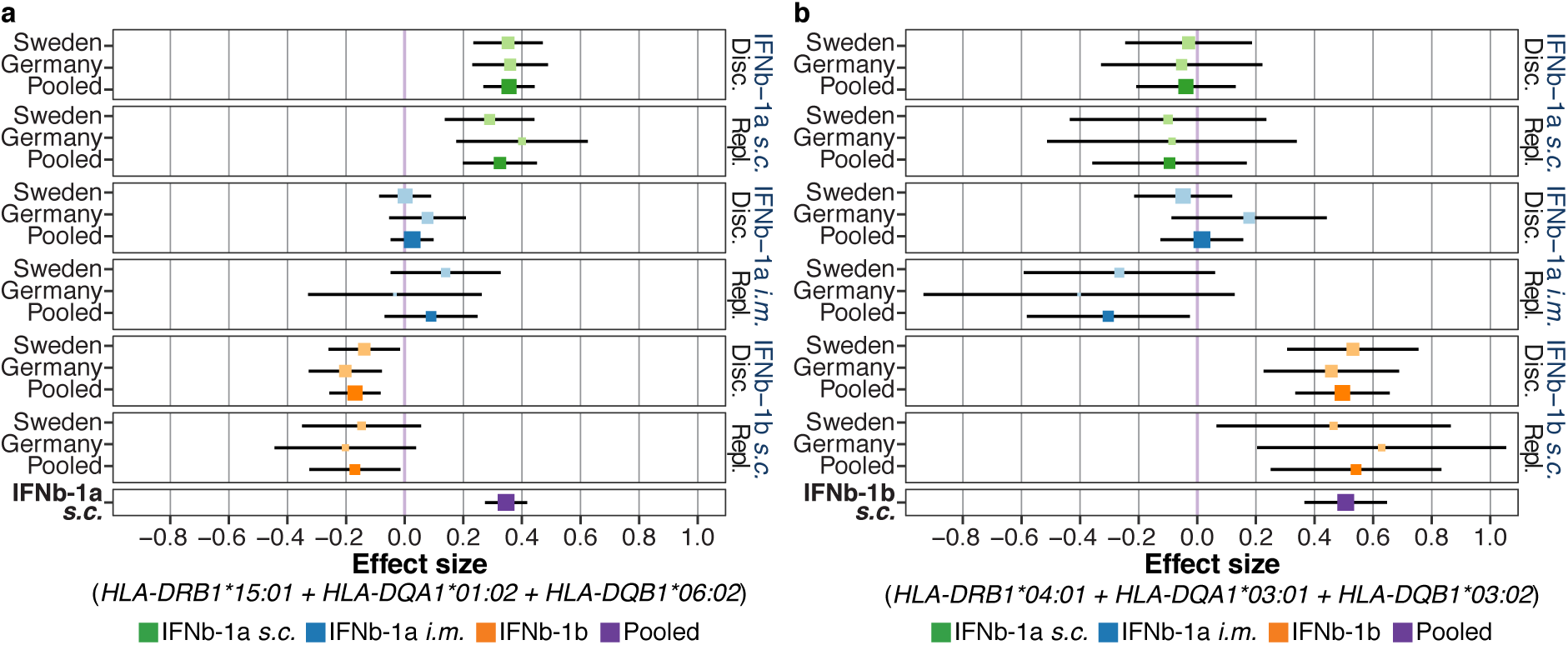
Treatment-specific *HLA* haplotypes. **a-b** The association of nADA titers for selected extended haplotypes showing strong treatment-specific effects. For association statistics, see Table 3 and S6 Table. Disc. = discovery, Repl. = replication. **a** The association of the *DR15-DQ6* haplotype with nADA titers is specific for IFNβ-1a *s.c*. **b** The association of the *DR4-DQ3* haplotype with nADA titers is specific for IFNβ-1b *s.c*.

### Treatment-specific GWAS: IFNβ-1a *s.c*

We also conducted treatment preparation-specific GWAS [Additional files 15 and 16]. In analyses of both nADA presence and titers in IFNβ-1a *s.c*.-treated patients, variant rs77278603 was genome-wide significant in the discovery stage and replicated (OR=3.55 (2.81-4.48), *p*=2.1×10^−26^; Table 2 [Additional files 8-10]). It maps downstream of *HLA-DRB5*. In the pooled analyses, two different but correlated SNPs upstream of *HLA-DQA1* were the most strongly associated signals, which were both significant eQTLs with an *HLA-DRB5* transcript [Additional file 12]. These two SNPs, rs9271700 for nADA presence and rs9271673 for nADA titers (Table 2 [Additional file 8]), and the top discovery-stage variant rs77278603 were in LD with allele *HLA-DRB1*15:01* (*r*^2^≥0.71). When conditioning the IFNβ-1a *s.c*.-associated risk *HLA* haplotype and alleles for rs77278603, none of them remained significant [Additional file 14]. The *DR15-DQ6* risk association from the *HLA* analysis thus likely corresponds to the association of rs77278603 and correlated SNPs.

For bADA levels in IFNβ-1a *s.c*.-treated patients, rs9281962 was the top-associated variant in both the discovery and pooled analysis (Table 2 [Additional file 8]), it was in very high LD with the nADA-associated SNPs rs9271700 and rs9271673 (*r*^2^≥0.93). Notably, none of the variants associated at genome-wide significance with ADA in IFNβ-1a *s.c*.-treated patients showed statistical support for an association in IFNβ-1b *s.c*.-treated patients with *p*<0.001 in the discovery stage. When analyzing both IFNβ-1a preparations together, results were highly similar to when analyzing IFNβ-1a *s.c*.-treated patients alone [Additional files 8 and 14].

### Treatment-specific GWAS: IFNβ-1b *s.c*

The SNP rs28366299 was significantly associated with nADA presence in IFNβ-1b *s.c*.-treated patients in both the discovery and pooled analysis (OR=3.56 (2.69-4.72), *p*=6.6×10^−19^; Table 2 [Additional file 8]). It maps upstream of *HLA-DRB1*, is correlated with *HLA-DQA1*03:01* (*r*^2^≥0.46) and an eQTL with an *HLA-DQA2* transcript [Additional file 12].

In conditional analyses of the IFNβ-1b *s.c*.-associated risk *HLA* haplotype and alleles for rs28366299, *HLA-DRB1*04:01* and *DR4-DQ3* remained significant [Additional file 14] and vice versa [Additional file 8], indicating that rs28366299 represents a signal independent of the *HLA* association. We analyzed this SNP in a study of nADA presence using a published independent study on 941 IFNβ-1b *s.c*.-treated patients [15], where it replicated robustly (OR 2.37 (1.81-3.08), one-sided *p*=9.88×10^−11^; meta-analysis with the present study: OR 2.87 (2.37-3.48), *p*=7.74×10^−27^).

The same variant was also associated with the nADA titers (Table 2 [Additional file 8]). SNP rs9272775, intronic in *HLA-DQA1* and correlated with rs28366299 (*r*^2^≥0.79), was the top variant in the pooled analysis of nADA titers. In the independent study [15], variant rs9272775 replicated with one-sided *p*=6.05×10^−17^ (meta-analysis *p*=7.62×10^−40^).

The analysis of bADA levels in IFNβ-1b *s.c*.-treated patients produced similar results (Table 2 [Additional file 8]). Here as well, rs9272775 was the top-associated SNP in the pooled analysis. In the discovery stage, rs78279385 (LD with rs9272775 *r*^2^≥0.88) was the variant showing the most robust support for an association. None of the variants associated at genome-wide significance with ADA in IFNβ-1b *s.c*.-treated patients showed statistical support for an association in IFNβ-1a *s.c*.-treated patients with *p*<0.009 in the discovery stage.

### Treatment-specific GWAS: Stepwise conditional analyses

In stepwise conditional regression analyses in the pooled dataset, none but the respective top-associated variants were significantly associated with IFNβ-1a *s.c*.-induced ADA [Additional file 11]. For IFNβ-1b *s.c*., rs559242105, in LD with *HLA-DPB1*03:01* (*r*^2^≥0.78), was identified as a secondary signal for nADA presence and titers when conditioning for the respective top SNP. In conditional analyses, this insertion was independent of *HLA-DRB1*04:01* (nADA titers without *HLA: p*=5.2×10^−10^, conditioned with *HLA:* 1.1×10^−08^). Possibly, the conditional association of rs559242105 corresponds to a secondary protective signal by *HLA-DPB1*03:01* (conditional model of *HLA-DPB1*03:01* OR=0.49 (0.35-0.69), *p*=4.7×10^−05^ [Additional file 11]).

For bADA levels, *HLA-DRB1*04:01* was the top association, followed by rs17205731 as a secondary signal. The two secondary variants rs559242105 and rs17205731 are not in LD with each other and both map to *HLA*-associated loci further downstream than the primary association signal [Additional file 9].

### Analysis of candidate variants

Association results of previously published variants associated with ADA are listed in [Additional file 17]. SNP rs9272105, mapping to the MHC region and previously identified in a study conducted on a subset of the patients analyzed here [14], was significantly associated across treatment preparations in the present study. However, we found no support for an association of variant rs4961252 on chromosome 8, identified in the same study [14], which confirms a previously failed replication attempt [15]. Both variants previously identified in an independent study of IFNβ-1b *s.c*.-treated patients [15] replicated only in individuals treated with IFNβ-1b *s.c*.

Of the previously reported associations with *HLA* alleles, associations with the risk alleles *HLA-DQA1*02:01, -DRB1*04:01, *04:08, *07:01, *08:01, *15:01*, and *\*16:01*, and with the protective alleles *HLA-DQA1 *05:01*, *-DQB1*02:01, -DRB1*03:01*, and *\*04:04*, replicated in at least one treatment preparation and ADA measurement when correcting for 20 tested variants [Additional file 17]. Notably, *HLA-DRB1*04:08* was only associated with nADA levels and nADA titers but not with nADA presence.

### Prediction of ADA

We predicted the occurrence of nADA in the replication dataset using the genetic models derived in the discovery dataset. For each treatment preparation, we analyzed eight polygenic risk scores (PRS), the top single GWAS variant, and the top *HLA* allele from the discovery stage (Fig 3a-b, [Additional files 18 and 19]). Based on the AUC, the best predictions were achieved in models either featuring only the top variant or by PRS consisting of variants showing strong support for an association (Fig 3). We thus did not observe evidence for a highly polygenic inheritance of nADA development. Interestingly, only treatment-specific predictions were significant; IFNβ-1a *s.c*.-specific models could not predict nADA in IFNβ-1b *s.c*.-treated patients and vice versa (Fig 3c [Additional file 20]). Prediction models containing either the top PRS or SNP showed distinctly increased AUCs, Nagelkerke’s pseudo-*R*^2^, sensitivities, and specificities over models containing just the covariates (Table 4).

**Fig 3:**
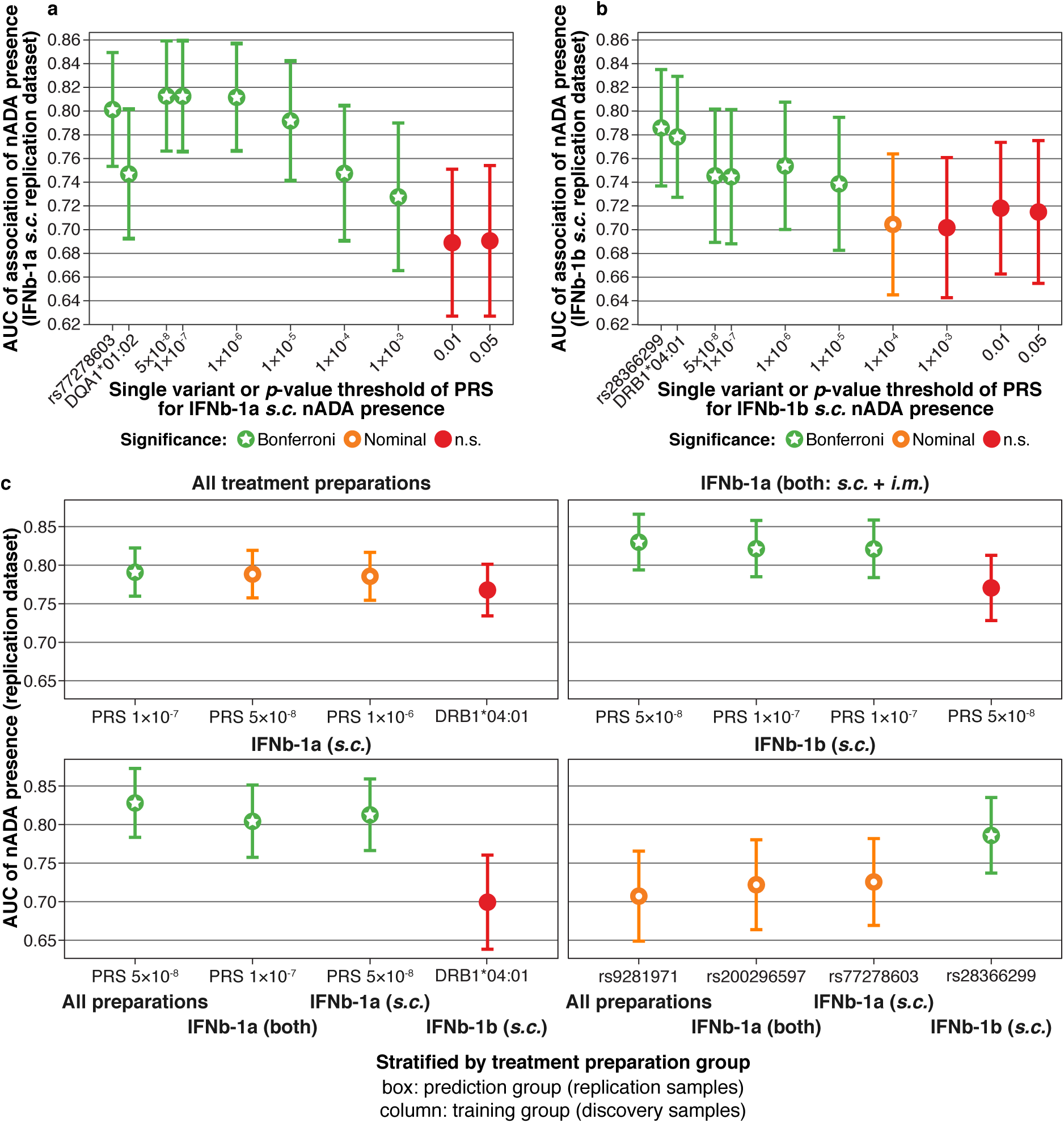
Prediction of nADA. Treatment-specific prediction of the presence of nADA in the replication data. Eight PRS, the top single GWAS variant, and the top *HLA* allele from the discovery stage were analyzed, with sex, age, treatment preparation and duration, titration site, and ancestry components as covariates. The plots show the area under the receiver operating characteristic curve (AUC) and its 95 % confidence interval (CI) calculated using bootstrapping. Bonferroni = significant after Bonferroni correction for multiple testing; nominal = nominally significant (*p*<0.05); n.s. = not significant. **a-b** AUC of all ten prediction models for **a** IFNβ-1a *s.c*. and **b** IFNβ-1b *s.c*.; Bonferroni correction for ten tests. **c** The model with the highest AUC for each treatment preparation, Bonferroni correction for 160 tests (α=3.13×10^−4^). Boxes show the prediction groups (replication data) and columns within each box the training data groups (discovery data).

**Table 4:** Treatment-specific prediction of the presence of nADA in the replication data. **Table 4:** Predictors in the model without genetics: sex, age, treatment duration, and titration site. The genetic models contained the same base model plus the indicated genetic factors and ancestry components. The top models are indicated in bold font. OR = odds ratio, CI = confidence interval, *p* = *p*-value of the genetic component, AUC = area under the receiver operating characteristic curve, *R^2^* = Nagelkerke pseudo-*R*^2^; KI = Karolinska Institutet, Sweden; TUM = Technical University of Munich, Germany.a

| Preparation | Model | Cohort | OR | 95% CI | *p* | AUC | *R*<sup>2</sup> | Sensitivity | Specificity |
| --- | --- | --- | --- | --- | --- | --- | --- | --- | --- |
| IFNβ-1a s.c. | Without genetics | KI |  |  |  | 0.65 | 0.08 | 0.66 | 0.52 |
| IFNβ-1a s.c. | Without genetics | TUM |  |  |  | 0.60 | 0.06 | 0.71 | 0.42 |
| IFNβ-1a s.c. | PRS nADA presence $5 \times 10^{-08}$ | KI | 3.89 | 2.35-6.45 | $1.44 \times 10^{-07}$ | 0.85 | 0.42 | 0.78 | 0.78 |
| IFNβ-1a s.c. | PRS nADA presence $5 \times 10^{-08}$ | TUM | 2.56 | 1.56-4.21 | $2.11 \times 10^{-04}$ | 0.76 | 0.24 | 0.68 | 0.65 |
| IFNβ-1a s.c. | **PRS top 30% vs. bottom 30%** | **KI** | **73.86** | **11.77-463.61** | **$4.42 \times 10^{-06}$** | **0.91** | **0.59** | **0.78** | **0.90** |
| IFNβ-1a s.c. | **PRS top 30% vs. bottom 30%** | **TUM** | **13.78** | **3.00-63.28** | **$7.45 \times 10^{-04}$** | **0.83** | **0.38** | **0.80** | **0.76** |
| IFNβ-1a s.c. | SNP rs77278603 additive coding | KI | 4.49 | 2.41-8.36 | $2.14 \times 10^{-06}$ | 0.82 | 0.36 | 0.74 | 0.74 |
| IFNβ-1a s.c. | SNP rs77278603 additive coding | TUM | 3.88 | 1.78-8.47 | $6.67 \times 10^{-04}$ | 0.73 | 0.21 | 0.63 | 0.74 |
| IFNβ-1a s.c. | SNP rs77278603-A dominant coding | KI | 9.16 | 2.48-33.79 | $8.79 \times 10^{-04}$ | 0.78 | 0.31 | 0.57 | 0.76 |
| IFNβ-1a s.c. | SNP rs77278603-A dominant coding | TUM | 3.85 | 1.10-13.49 | $3.51 \times 10^{-02}$ | 0.72 | 0.20 | 0.56 | 0.68 |
| IFNβ-1b s.c. | Without genetics | KI |  |  |  | 0.70 | 0.15 | 0.48 | 0.82 |
| IFNβ-1b s.c. | Without genetics | TUM |  |  |  | 0.58 | 0.02 | 0.82 | 0.20 |
| IFNβ-1b s.c. | PRS nADA presence $1 \times 10^{-06}$ | KI | 2.40 | 1.45-3.97 | $6.46 \times 10^{-04}$ | 0.78 | 0.33 | 0.57 | 0.85 |
| IFNβ-1b s.c. | PRS nADA presence $1 \times 10^{-06}$ | TUM | 2.15 | 1.43-3.23 | $2.28 \times 10^{-04}$ | 0.73 | 0.22 | 0.73 | 0.58 |
| IFNβ-1b s.c. | PRS top 30% vs. bottom 30% | KI | 10.16 | 2.30-44.95 | $2.25 \times 10^{-03}$ | 0.83 | 0.46 | 0.58 | 0.87 |
| IFNβ-1b s.c. | PRS top 30% vs. bottom 30% | TUM | 5.97 | 2.03-17.52 | $1.14 \times 10^{-03}$ | 0.78 | 0.33 | 0.69 | 0.75 |
| IFNβ-1b s.c. | SNP rs28366299 additive coding | KI | 4.51 | 1.72-11.80 | $2.14 \times 10^{-03}$ | 0.77 | 0.31 | 0.57 | 0.82 |
| IFNβ-1b s.c. | SNP rs28366299 additive coding | TUM | 6.91 | 3.18-15.03 | $1.07 \times 10^{-06}$ | 0.79 | 0.32 | 0.74 | 0.61 |
| IFNβ-1b s.c. | **SNP rs28366299-A dominant coding** | **KI** | **9.78** | **2.68-35.74** | **$5.62 \times 10^{-04}$** | **0.83** | **0.40** | **0.62** | **0.83** |
| IFNβ-1b s.c. | **SNP rs28366299-A dominant coding** | **TUM** | **7.56** | **3.01-19.02** | **$1.71 \times 10^{-05}$** | **0.80** | **0.33** | **0.77** | **0.57** |

Finally, patients with a high and low genetic risk burden were contrasted [46]. To this end, patients within the upper 30% of the top-associated PRS were compared to the patients within the lower 30%. This specific threshold was set to allow for a large enough sample size in the replication dataset for the stable convergence of regression models when conducting cross-validation. In addition, the respective top SNP was analyzed using dominant coding, thereby comparing no copy of the risk allele to any copy. Here the best prediction was achieved for Swedish patients treated with IFNβ-1a *s.c*., with an AUC=0.91 (CI=0.85-0.95), a pseudo-*R*^2^=0.59, sensitivity=0.78, and specificity=0.90. Patients with the top 30% of genetic risk had, compared to patients in the bottom 30%, an OR=73.9 (CI=11.8-463.6, *p*=4.4×10^−06^) of developing nADA.

## Discussion

Several studies have previously assessed genetic risk factors for ADA. They were limited by much smaller sample sizes, only analyzed a single cohort, focused mostly on *HLA* alleles, and did not consistently assess ADA with sensitive and validated methods. The latter is also reflected in the increased number of nADA-positive samples in the measurements conducted for the present study, compared to previous results [Additional files 1 and 3]. Most importantly, the existing studies neither reached a consensus on genetic risk factors nor could they delineate a robust prediction model for ADA. To our knowledge, the present study constitutes the most extensive genetic characterization of ADA risk to date, is the first systematic comparison of the genetics of different ADA types and includes the first genetic prediction model for ADA against IFNβ.

### Previously reported genetic risk factors

All genetic variants robustly associated with ADA in the present study map to the MHC region and are linked to the expression of *HLA* genes or amino acid changes in the peptide-binding groove of HLA molecules. The only previously published non-MHC risk variant [14] replicated neither in an earlier [15] nor in the present study.

Previous studies prioritized sixteen different *HLA* alleles as potentially associated with nADA presence, nADA titers, or bADA levels [10–13, 15, 16]. Of these, eleven replicated in our study for any treatment, and five did not. Importantly, the present study does not constitute a formal replication for many of the candidate *HLA* alleles because of the extensive sample overlap with previous Swedish and German studies. The variants that did not replicate had low frequencies, with a maximum AF=0.06, and showed only weak support in the original studies. Notably, some previous studies analyzed highly correlated alleles as if they were independent variants or did not correct for multiple testing. Moreover, Núñez *et al*. analyzed Spanish patients [16], which differ in their allele frequencies and linkage patterns from the individuals studied here. Next to population-specific effects, joint analyses of samples receiving different treatment preparations constituted a source of heterogeneity in several previous studies. In our comprehensive analyses, we could now consolidate the many *HLA* alleles published in previous studies into few treatment-specific haplotypes

### Treatment preparation-specific risk

In the analyses across treatment preparations, the associations of the replicated GWAS SNPs were supported in all analyzed treatment groups [Additional file 10]. Nevertheless, we observed lower *p*-values and larger effect sizes in the preparation-specific analyses than in the models combining patients across treatments. The combination of patients receiving different treatment preparations thus created heterogeneity that decreased statistical power. This hypothesis was further supported in stepwise conditional analyses. Here, we observed more evidence for the existence of independent risk loci in analyses across preparations than was the case in treatment-specific analyses. Likely, such presumably independent loci in the combined analysis reflect treatment preparation-specific effects. These findings thus argue in favor of conducting treatment-specific over cross-treatment analyses. In future studies of ADA against biopharmaceuticals, analyses of preparation-specific risk factors should, therefore, be prioritized.

Although differences in the antigenic potential of the various IFNβ preparations are known [1], the extent of preparation-specific genetic risk observed in the present study is striking (Fig. 2). There are several plausible explanations for why the preparations might be processed differently by the immune system. While the amino acid sequence of IFNβ-1a is identical to natural human IFNβ, IFNβ-1b diverges at two positions: IFNβ-1b lacks the N-terminal methionine, and a cysteine at position 17 is substituted by serine. Furthermore, the products are raised in different cell types, prokaryotic *E. coli* and eukaryotic Chinese hamster ovary cells, leading to different post-translational modifications, especially glycosylation [1].

Lack of glycosylation facilitates the formation of protein aggregates, increasing the immunogenicity of IFNβ-1b [1, 2]. Previous research demonstrated that, among the three preparations analyzed in the present study, IFNβ-1b shows the highest tendency to aggregate [47]. IFNβ-1a *i.m*., which does not contain human serum albumin, forms the fewest aggregates and shows the lowest rate of ADA. In contrast to IFNβ-1b *s.c*., aggregates in IFNβ-1a *s.c*. preparations are mainly formed by human serum albumin [47]. Differences in IFNβ protein aggregation might, in addition to increased presentation of peptides by dendritic cells and, thus, increased T cell activation [48, 49], also contribute to the diversification of genetic risk factors. When taken up by antigen-presenting cells, e.g., dendritic cells, IFNβ oligomers are likely degraded differently from monomers. Such differences in processing could produce diverse peptides, which may be presented by different MHC class II molecules [48].

Post-translational modifications not only affect aggregate formation but, together with differences in the amino acid sequences, also alter the biochemical properties of IFNβ-1a and IFNβ-1b. Thereby, both post-translational modifications and differences in the amino acid sequence may contribute to the preparation-specific associations with *HLA* alleles [4]. For example, altering epitopes by glycosylation strongly affects antigen recognition [50]. Possibly, glycosylated IFNβ-1a peptides are thus preferentially recognized by different MHC peptide-binding grooves than IFNβ-lb-derived epitopes are. Similarly, also the amino acid changes may alter the binding of IFNβ peptides to MHC molecules and T cell recognition.

Additional factors in the processing of treatment preparations can influence how the immune system recognizes them. Spontaneously occurring modifications like deamidation, oxidation, and glycation alter the surface and chemical properties of proteins. These modifications even diverge between preparations sharing the identical amino acid sequence, e.g., IFNβ-1a *s.c*. and *i.m*., by differential production, processing, or storing of the biopharmaceuticals [3]. Other chemical alterations of amino acids like phosphorylation, PEGylation, methylation, or acetylation can be applied during the manufacturing of drugs, e.g., to alter their stability, and also change epitopes, leading to differential recognition by *HLA* alleles [51, 52]. Importantly, these modifications also happen after administration of the product *in vivo*, and glycosylation may well affect the likelihood of them taking place.

In summary, diverging post-translational modifications may contribute to the observed differences in preparation-specific genetic risk factors. Notably, the MHC class II peptide-binding groove is formed by heterodimers of two HLA proteins, likely contributing to the association of haplotypes spanning *HLA* α and β chain genes, like *HLA-DQA1* and *HLA-DQB1*, with IFNβ ADA.

However, it is unlikely that preparation-specific risk can entirely be attributed to genetic factors. For example, the dosage and injection frequency of preparations may affect the likelihood of developing ADA, independently of genetic risk. Nevertheless, most patients develop ADA within the first months of IFNβ treatment [53], arguing against pronounced long-term dosage-specific effects and underlining the importance of genetic risk.

Next to having to rely on imputed *HLA* alleles, the low number of available patients that developed ADA under treatment with IFNβ-1a *i.m*., rendering IFNβ-1a i.m.-specific analyses unfeasible, constitutes a limitation of the present study. We expect genetic risk factors for IFNβ-1a *i.m*.-induced ADA to exist, but whether these are independent of IFNβ-1a *s.c*.-associated risk remains to be shown.

### The complexity of the genetic risk landscape

Using conditional analyses, we did not find evidence for more than one genetic risk locus for IFNβ-1a *s.c*.-induced ADA. Results from previous studies can thus, at least for Swedish and German patients, be consolidated to the extended haplotype *DR15-DQ6*. In the present dataset, it is impossible to assess whether the combined *DR15-DQ6* haplotype constitutes the real risk factor for IFNβ-1a-*s.c*. or whether any of the single alleles *HLA-DQB1*06:02* or *HLA-DQA1*01:02* convey this risk, with the haplotype showing an association merely because of LD. *DR15-DQ6* (MAF_KI_=0.34, MAF_TUM_=0.29) is less common than the two single alleles, especially compared to *HLA-DQA1 *01:02* (MAF_KI_=0.42, MAF_TUM_=0.36). Because statistical power is dependent on the AF, the slightly lower statistical support for the association of *DR15-DQ6*, compared to the single alleles, likely reflects these differences in AF and power. We thus hypothesize that the combined haplotype *DR15-DQ6* constitutes the primary signal. Nevertheless, such fine-mapping and the differentiation between the correlated alleles is irrelevant for risk predictions. Because of the strong correlation of alleles observed within the extended haplotype, any of these alleles can reliably be used as a proxy for the others in prediction models.

Similarly, conditional analyses support the association of the extended haplotype *DR3-DQ2* as the primary protective genetic signal for IFNβ-1a *s.c*., without evidence for secondary signals. However, in the present sample, the association of this haplotype cannot be separated from *HLA-DQB1*02:01*. By contrast, genetic risk for IFNβ-1b *s.c*.-induced ADA appears to be more complicated. The association of the haplotype *DR4-DQ3* could not fully explain the signal of its allele *HLA-DRB1*04:01*. Moreover, we found evidence for a secondary signal in stepwise conditional regression analyses. Notably, the prediction models for IFNβ-1b *s.c*. did not perform as well as the prediction for IFNβ-1a *s.c*.-induced ADA did, which possibly reflects this more complex risk landscape. To truly unravel an additional potential polygenic contribution to ADA risk, the current study still lacked the sample size necessary for reliably detecting polygenic variants with their expected small effect sizes [54, 55].

### Population-specific risk differences

Alleles from all three associated haplotypes, *HLA-DRB1*15:01, HLA-DRB1*04:01*, and the protective *HLA-DRB1*03:01*, are concurrent risk factors for MS [42]. The unfortunate coincidence that *HLA-DRB1 *15:01* and *HLA-DRB1*04:01* are enriched among MS patients and also constitute ADA risk factors likely contributes to the high incidence of IFNβ ADA among MS patients.

Interestingly, these alleles also show substantial population-specific differences [56]: The risk allele *HLA-DRB1 *15:01* is especially frequent in individuals with ancestry from parts of Northwestern, Northern, Central, and Eastern Europe (e.g., Northern Spain: 16.7-32.1%, Germany: 12.9-17.2%). At the same time, this allele is less frequent in Southern Europe and for Ashkenazi, Southern Hispanic, and African ancestry (e.g., Italy: 5.6-6.4%, Southwestern Spain: 5.2-8.6%). The risk allele *HLA-DRB1 *04:01* is more frequent in parts of Northwestern, Northern, and Central Europe (e.g., England: 12.4-13.5%, Denmark: 17.6%) than in Southern Europe and most other ancestries (e.g., Italy: 1.7-4.1%, Spain: 2.0-3.8%).

While the frequencies of both risk alleles for IFNβ-1a *s.c.-*and IFNβ-1b *s.c*.-induced ADA thus roughly decrease along a North-South gradient within Europe, their relative frequencies differ sharply in some ancestries [Additional file 21]. For example, in Northern Spain, the major genetic risk factor for IFNβ-1a *s.c*.-specific ADA occurs >8 times more often than the one for IFNβ-1b *s.c*.-induced ADA.

Such substantial, population-specific differences in risk allele frequencies likely exist for ADA against any biopharmaceutical. If genetic risk factors for a biopharmaceutical are known, and personalized genotyping data for patients are not available, recommendations for the choice of a specific treatment preparation could thus be made on a population level.

### Genetic factors contributing to nADA titers

We did not observe a significant genetic component for nADA titers in nADA-positive patients. Moreover, the same or highly correlated risk factors contributed to the presence of nADA and the magnitude of nADA titers and bADA levels. The heritability of nADA, explained by common variants, after correction for confounders like treatment preparation and duration, sex, and age, was very high, with 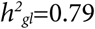 on a liability scale. This result underlines the importance of genetic factors in the occurrence of IFNβ ADA. Although nADA titers need to cross a threshold to become functionally relevant, the genetic risk surprisingly appears to mainly influence the likelihood of developing ADA and less the absolute titers. However, the association of the candidate variant *HLA-DRB1 *04:08* was only significant for bADA levels and nADA titers but not for nADA presence. This finding indicates that genetic factors influencing the amount of ADA may nevertheless exist.

### Prediction of ADA

We could predict nADA especially well in Swedish patients receiving IFNβ-1a *s.c*. Our results indicate that, compared to IFNβ-1b *s.c*., genetic risk for IFNβ-1a *s.c*.-induced ADA is more dominated by a single locus. In addition, overall, more patients receiving IFNβ-1a *s.c*. than IFNβ-1b *s.c*. were analyzed (1,145 vs. 1,010). Both factors likely contributed to better prediction models and performance in IFNβ-1a *s.c*.-treated patients. The top-associated ADA risk SNP was more frequent in Swedish patients than in German ones (43% *vs*. 40%), and the Swedish sample contained more patients treated with IFNβ-1a *s.c*. (Sweden: 590, Germany: 558), of whom more were nADA-positive (34.6% *vs*. 33.7%). Although these individual differences were small, they may have contributed to prediction models performing better in the Swedish dataset.

Contrasting the samples in the top and bottom percentiles of polygenic risk score distributions is a common practice to compare individuals carrying a high genetic risk burden to the ones not at risk [46]. The sensitivity and specificity reached in the comparison of individuals in the top 30% nADA risk group compared to the bottom 30% (0.78 and 0.90, respectively, in Swedish IFNβ-1a *s.c*. patients) may still not be sufficient for a routine clinical test. However, these prediction models could certainly be optimized by the inclusion of additional predictive factors, e.g., body mass index [57], not available in the present retrospective setting. The significant predictive improvement of the genetic risk model compared to a model containing only demographic and clinical variables (Table 4) underlines the importance of incorporating genetics in prediction models for ADA. The high odds of Swedish patients at genetic risk for nADA (OR=73.9) support the use of genetic stratification as a personalized medicine tool – patients at high genetic risk should either switch to a different drug or be monitored more closely, as suggested for other conditions [58].

## Conclusions

We have conducted a comprehensive characterization of genetic risk for IFNβ-induced ADA, consolidating previous research. Next to treatment-specific risk factors, we described ancestry-specific effects relevant for treatment choice in specific populations. Our robust prediction models could be employed for personalized medicine, guiding treatment recommendations, and efficient nADA testing regimes. Importantly, our study can serve as a blueprint for the analysis of genetic factors influencing ADA against other biopharmaceuticals and in the context of further diseases.

## Data Availability

The datasets generated and analyzed during the current study are available from the corresponding authors on reasonable request.

## List of abbreviations

ADA: anti-drug antibodies
AF: allele frequency
AUC: area under the receiver operating characteristic curve
bp: base pairs
bADA: binding ADA
CI: 95 % confidence interval
eQTL: expression quantitative trait locus
GWAS: genome-wide association study
IFNβ: Interferon β
*i.m*.: intramuscular
HLA: human leukocyte antigen
kbp: kilo base pairs
KI: Karolinska Institutet Stockholm Sweden
LD: linkage disequilibrium
MAD: median absolute deviation
MHC: major histocompatibility complex
MAF: minor allele frequency
MDS: multidimensional scaling
MS: multiple sclerosis
nADA: neutralizing ADA
OR: odds ratio
*s.c*.: subcutaneous
PRS: polygenic risk scores
SD: standard deviation
SE: standard error
SNP: single nucleotide polymorphism
TUM: Technical University of Munich, Germany

## Declarations

### Ethics approval and consent to participate

The ethics committees of the *Regionala etikprövningsnämnden i Stockholm* for Karolinska Institutet, Stockholm, Sweden, and of the Klinikum rechts der Isar, Technical University of Munich, Germany, approved the study, and all participants provided written informed consent.

### Consent for publication

Not applicable

## Author Contributions

TFMA drafted the conceptual study design, administered the project, curated data, devised the methodology for data analysis, conducted and interpreted the formal statistical data analysis, generated the data visualizations, and has prepared and revised the original manuscript draft. JL drafted the conceptual study design, curated and provided data on and investigated the Swedish patients, and revised the manuscript. DM drafted the conceptual study design, acquired funding, recruited and investigated the German patients, curated and provided their data, and revised the manuscript. MR curated data on and investigated the Swedish patients and revised the manuscript. CH curated data on and investigated the Swedish patients and reviewed the manuscript. VG performed the ELISA measurements of bADA levels and acquired data. MA contributed to nADA screening and titration, acquired data, and revised the manuscript. HH contributed to nADA screening and titration, acquired data, and revised the manuscript. LA recruited and acquired data on German patients and revised the manuscript. CG recruited and acquired data on German patients and revised the manuscript. BK recruited and acquired data on German patients and reviewed the manuscript BMM devised the methodology for data analysis, supervised statistical data analyses, and reviewed the manuscript. PEHJ conducted nADA screening and titration, acquired data, and reviewed the manuscript. FS contributed to nADA screening and titration and reviewed the manuscript. IK provided data on the Swedish patients. TO provided data on the Swedish patients and reviewed the manuscript MP acquired funding, administered the project, and reviewed the manuscript. SeS drafted the conceptual study design, acquired funding, administered the project, and revised the manuscript. FD drafted the conceptual study design, acquired funding, administered the project, devised the methodology for ADA measurement, conducted nADA screening and titration, acquired data, and revised the manuscript. AFH drafted the conceptual study design, acquired funding, administered and supervised the project, devised the methodology for ADA measurement, provided data on Swedish patients, and revised the manuscript. BH drafted the conceptual study design, acquired funding, supervised the project, provided data on German patients, and revised the manuscript.

## Acknowledgments

We are thankful for the excellent technical work preparing samples and measuring nADA by the technical staff, Birgit Kassow and Ulla Abildtrup, of DMSC, Department of Neurology, Rigshospitalet, University of Copenhagen, Denmark, Marlies Jank of the Department of Neurology, Medical University of Innsbruck, Austria, and Anna Mattsson, of Karolinska Institutet, Stockholm, Sweden. We thank Niek de Vries, Faculty of Medicine, University of Amsterdam, Netherlands, for helpful discussions on *HLA* haplotypes.

## Additional files

### Additional file 1.pdf

**Previous measurements and design of new ADA measurements**

Previous ADA measurements in the Swedish KI and German TUM cohorts per treatment preparation and distribution of samples for the new ADA measurements. For part of the TUM patients, only previous bADA measurements were available.

### Additional file 2.docx

**Additional details supporting the Methods section**

### Additional file 3.pdf

**New ADA measurements and design of the datasets for analyses**

New ADA measurements in the Swedish KI and German TUM cohorts per treatment preparation and assignments of samples into the discovery and replication datasets. In the discovery and replication datasets, the first number indicates nADA and the second number bADA measurements. The distinction into negative and positive patients was made using nADA measurements.

### Additional file 4.docx

**Visualization of population stratification**

### Additional file 5.png

**Manhattan plots of the GWAS on combined treatments**

Manhattan plots of the (**A-C**) discovery-stage and (**D-F**) pooled discovery + replication GWAS. The red line between –log_10_*p* =7 and –log_10_*p*=8 indicates genome-wide significance; the top genome-wide significant variant is labeled with a red diamond.

### Additional file 6.png

**Manhattan plots of the MHC region of the GWAS on combined treatments**

Manhattan plots of the (**A-C**) discovery-stage and (**D-F**) pooled discovery + replication GWAS, showing only the MHC region. The red line between –log_10_*p* =7 and –log_10_*p* =8 indicates genome-wide significance. For (**A-C**) discovery-stage plots, the prioritized variants are labeled with red diamonds, for (**DF**) pooled discovery + replication plots, the replicated variants are labeled with red diamonds, and the top pooled variant is labeled in magenta.

### Additional file 7.xlsx

**Genomic inflation factors for all GWAS**

Lambda = Median genomic inflation factor.

### Additional file 8.xlsx

**Table of the top GWAS associations**

Variants prioritized in the discovery GWAS (bold font if replicated) and top variants from the pooled analysis of discovery + replication data. All effect sizes are relative to the minor allele. Bp = base pairs, MAF = minor allele frequency, beta = regression effect size, SE = standard error, P = *p*-value, cond. = conditional analysis, R2 = linkage disequilibrium *r^2^*.

### Additional file 9.docx

**Regional association plots of the top GWAS variants**

Regional association plots of variants from the GWAS generated using LocusZoom v1.4 and the l000 Genomes l000G_Nov20l4 EUR reference panel [59]. The color of dots indicates LD with the lead variant (pink). Gray dots represent signals with missing LD *r^2^* values. If no LD information was present in the database on the top variant, LD with the variant showing the second-lowest *p*-value is indicated. The grey line indicates genome-wide significance. cM: centimorgan, chr: chromosome, Mb: mega base pairs.

### Additional file 10.docx

**Forest plots of the top GWAS variants and *HLA* alleles**

Green: IFNβ-1a *s.c*., blue: IFNβ-1a *i.m*., orange: IFNβ-1b *s.c*., magenta: pooled discovery-/replication-stage analyses. D. = discovery, R. = replication, P. = pooled discovery + replication.

### Additional file 11.xlsx

**Results from step-wise conditional analyses**

### Additional file 12.xlsx

**Results from eQTL analyses**

Summary statistics as downloaded from GTEx v8 (https://gtexportal.org/). Significance thresholds are shown in the column *Gene-level P threshold*.

### Additional file 13.xlsx

**Results from MAGMA gene set analyses**

FDR = 5 % false discovery rate.

### Additional file 14.xlsx

**Association statistics of all replicated *HLA* alleles**

AF = allele frequency, beta = regression effect size, SE = standard error, P = *p*-value, cond. = conditional analysis.

### Additional file 15.png

**Manhattan plots of the GWAS on patients treated with IFNβ-1a *s.c*. or IFNβ-1b *s.c***.

Manhattan plots of the (**A-C, G-I**) discovery-stage and (**D-F, J-L**) pooled discovery + replication GWAS. The red line between –log_10_*p*=7 and –log_10_*p*=8 indicates genome-wide significance; the top genome-wide significant variant is labeled with a red diamond. (**A-F**) shows analyses for IFNβ-1a *s.c*., (**GL**) for IFNβ-1b *s.c*.

### Additional file 16.png

**Manhattan plots of the MHC region of the GWAS on patients treated with IFNβ-1a *s.c*. or IFNβ-1b *s.c***.

Manhattan plots of the (**A-C, G-I**) discovery-stage and (**D-F, J-L**) pooled discovery + replication GWAS, showing only the MHC region. The red line between –log_10_*p*=7 and –log_10_*p*=8 indicates genome-wide significance. For (**A-C, G-I**) discovery-stage plots, the prioritized variants are labeled with red diamonds, for (**D-F, J-L**) pooled discovery + replication plots, the replicated variants are labeled with red diamonds, and the top variant from the pooled analysis is labeled in magenta. (**A-F**) shows analyses for IFNβ-1a *s.c*., (**G-L**) for IFNβ-1b *s.c*.

### Additional file 17.xlsx

**Results from candidate SNP and *HLA* allele analyses**

Variants and alleles are labeled in bold font if they replicated in the present study. All effect sizes are relative to the minor allele. Bp = base pairs, MAF = minor allele frequency, AF = allele frequency, beta = regression effect size, SE = standard error, P = *p*-value, cond. = conditional analysis.

### Additional file 18.xlsx

**Treatment preparation-specific prediction of the presence of nADA in the replication data**

Eight PRS, the top single GWAS variant, and the top *HLA* allele from the discovery stage were analyzed in the replication data using the covariates sex, age, treatment preparation, treatment duration, titration site, and eight ancestry components. Upper table: Prediction of the presence of nADA in IFNβ-1a *s.c.-*treated patients from the replication data using all ten prediction models based on analyses for IFNβ-1a *s.c*. in the discovery data. Lower table: Prediction of the presence of nADA in IFNβ-1b *s.c*.-treated patients from the replication data using all ten prediction models based on analyses for IFNβ-1b *s.c*. in the discovery data. Beta = regression effect size, SE = standard error, P = *p*-value.

### Additional file 19.docx

**Treatment preparation-specific prediction of the presence of nADA in the replication data: performance of single models**

Eight PRS, the top single GWAS variant, and the top HLA allele from the discovery stage. Covariates: sex, age, treatment preparation, treatment duration, titration site, and ancestry components. The plots show the area under the receiver operating characteristic curve (AUC) and its 95 % confidence interval (CI). Bonferroni = significant after Bonferroni correction for multiple testing; nominal = nominally significant (*p*<0.05); n.s. = not significant

### Additional file 20.docx

**Treatment preparation-specific prediction of the presence of nADA in the replication data: comparison of top models**

For each top model, the plots show either the AUC and its 95 % CI or Nagelkerke’s pseudo-R^2^ and its 95 % CI. Boxes show the prediction groups (replication data) and columns within each box the training data groups (discovery data). Bonferroni = significant after Bonferroni correction for multiple testing; nominal = nominally significant (*p*<0.05); n.s. = not significant.

### Additional file 21.pdf

**Comparison of allele frequencies for *HLA-DRB1 *15:01* and *HLA-DRB1 *04:01***

The allele frequencies (AF) were queried from allelefrequencies.net on April 27^th^ 2020 [56]. All four-digit European Silver and Gold populations with data on both *HLA-DRB1 *15:01* and *HLA-DRB1 *04:01* were used. Populations with an AF below the average for *HLA-DRB1*15:01* and above the average for *HLA-DRB1*04:01* are colored in green. Populations with an AF above the average for *HLA-DRB1 *15:01* and below the average for *HLA-DRB1*04:01* are colored in magenta. In addition, the European populations with the highest or lowest AF for the respective allele as well as the largest German population are shown in grey. No Swedish population besides the Sami were available.

